# mRNA-1273 vaccine effectiveness against symptomatic SARS-CoV-2 infection and COVID-19-related hospitalization in children aged 6 months to 5 years

**DOI:** 10.1101/2023.06.27.23291933

**Authors:** Mary Aglipay, Jonathon Maguire, Sarah Swayze, Ashleigh Tuite, Muhammad Mamdani, Charles Keown-Stoneman, Catherine Birken, Jeffrey C Kwong

**Author notes:** **Correspondence to:** Jeffrey C Kwong, Senior Scientist, ICES, V1 06, 2075 Bayview Avenue, Toronto, Ontario, Canada, M4N 3M5, (or @DrJeffKwong on Twitter).

## Abstract

**Importance:** Data on mRNA-1273 (Moderna) vaccine effectiveness in children aged 6 months to 5 years are limited.

**Objective:** To assess mRNA-1273 vaccine effectiveness against symptomatic SARS-CoV-2 infection and COVID-19-related hospitalization among children aged 6 months to 5 years.

**Design, Setting, and Participants:** A test-negative study using linked health administrative data in Ontario, Canada. Participants included symptomatic children aged 6 months to 5 years who were tested by RT-PCR.

**Exposures:** mRNA-1273 vaccination.

**Main Outcomes and Measures:** Symptomatic SARS-CoV-2 infection and COVID-19-related hospitalization.

**Results:** We included 3467 test-negative controls and 572 test-positive cases. Receipt of mRNA-1273 was associated with reduced symptomatic SARS-CoV-2 infection (VE=90%; 95%CI: 53, 99%) and COVID-19-related hospitalization (VE=82%; 95%CI: 4, 99%) ≥7 days after the second dose.

**Conclusions and Relevance:** Our findings suggest mRNA-1273 vaccine effectiveness is initially strong against symptomatic SARS-CoV-2 infection and hospitalization in children aged 6 months to 5 years. Further research is needed to understand long-term effectiveness and the need for booster doses.

## INTRODUCTION

Vaccination has been identified as an important strategy to mitigate the impact of SARS-CoV-2. In the Canadian province of Ontario, the mRNA-1273 (Moderna) COVID-19 vaccine became available for children aged 6 months to 5 years on July 28, 2022, with a recommended dosing interval between the 2 doses of at least 8 weeks.

Clinical trial data have shown the efficacy of mRNA-1273 against symptomatic SARS-CoV-2 infection to be 36.8% for participants aged 2 to 5 years and 50.6% for participants aged 6 months to 23 months during a period of omicron dominance.^1^

Real-world data on COVID-19 vaccine effectiveness in children aged 6 months to 5 years are limited. These data are needed to inform public health policies and family decision-making around COVID-19 vaccination. We sought to evaluate mRNA-1273 vaccine effectiveness (VE) against symptomatic SARS-CoV-2 infection and COVID-19-related hospitalization among children aged 6 months to 5 years.

## METHODS

We conducted a retrospective test-negative study to evaluate the effectiveness of the mRNA-1273 vaccine in Ontario against symptomatic SARS-CoV-2 infection and COVID-19-related hospitalization using linked population-based databases. These datasets were linked using unique encoded identifiers and analyzed at ICES. We included community-dwelling children aged 6 months to 5 years as of July 28, 2022 who underwent testing for SARS-CoV-2 by real-time polymerase chain reaction (RT-PCR) and had documented signs or symptoms consistent with SARS-CoV-2 infection (see Supplementary Text).

Testing information and symptoms were captured through the Ontario Laboratories Information System. The study period was from July 28, 2022 to December 31, 2022, an omicron-dominant phase of the pandemic (>97% of all cases).^2,3^ In response to large volumes, diagnostic RT-PCR testing was restricted to specific populations during this time including hospitalized patients, patients seen in emergency departments, outpatients for whom COVID-19 treatment was being considered, household members of patient-facing healthcare workers, and symptomatic students who received a RT-PCR self-collection kit through their school.^4^

We excluded children who were immunocompromised,^4^ those without provincial health insurance, those missing birthdate, sex, or postal code information, and those who received the BNT162b2 (Pfizer) vaccine, a non-Health Canada authorized vaccine, or a dose of any COVID-19 vaccine prior to July 28, 2022. We also excluded children who were tested <14 days after their first vaccine dose, and children who had another positive test ≤90 days prior to specimen collection.

The primary outcome was a positive test for symptomatic SARS-CoV-2 infection, ascertained through RT-PCR. Children who tested positive for symptomatic SARS-CoV-2 infection at least once during the study period were considered cases. For children with more than one positive test, we used the first test. For children with multiple negative tests, we randomly selected a negative test for the analysis. The index date was the date of specimen collection.

The secondary outcome was hospitalization due to, or partially due to, COVID-19. According to the guidelines for data entry in the public health COVID-19 surveillance database, information about hospitalization should only be included for those cases where the patient was admitted to hospital for COVID-19 treatment or if their hospital stay was prolonged as a result of COVID-19. The primary exposure was receipt of the mRNA-1273 vaccine, as recorded in COVaxON, a centralized immunization information system that includes comprehensive information on all COVID-19 vaccination events in Ontario.^4,5^

Covariates were chosen a priori and evaluated as potential confounders (see Supplementary Table 2) based on their known relationships with SARS-CoV-2 infection or severity and receipt of a COVID-19 vaccine. These variables included (and are detailed elsewhere^6^) age, sex, public health unit region, comorbidities, influenza vaccination status (ever versus never), number of physician visits during the past year, mother’s healthcare worker status, and neighbourhood-level data from the 2016 Census on household income quintile, household density quintile, visible minority quintile, and non-health essential worker quintile.

We used a test-negative design to estimate VE of one or two doses of the mRNA-1273 against symptomatic laboratory-confirmed SARS-CoV-2 infection or COVID-19-related hospitalization. VE was defined as 1 minus the odds ratio of vaccination among cases compared to controls.^7^ We used multivariable logistic regression to adjust for potential confounding factors. Using interaction terms in multivariable logistic regression models and comparing models with and without interaction terms using likelihood ratio tests (LRT), we examined effect modification by age (6 months to < 2 years and 2 years to 5 years) and positive SARS-CoV-2 RT-PCR test ≥ 90 days before index date. All analyses were conducted using R version 4.1.3 (R Foundation for Statistical Computing, Vienna, Austria) and SAS version 9.4 software (SAS Institute, Cary, NC)

## RESULTS

From July 28, 2022 to December 31, 2022, 4039 children were included in the analysis, with 572 (14.2%) symptomatic test-positive cases and 3467 (85.8%) symptomatic test-negative controls (Figure 1). In our sample, test-positive cases were more likely to be less than 1 year of age (SMD=0.30), more likely to live in areas with the highest household density (SMD=0.20), and more likely to live in areas other than the middle quintile for visible minority (Table 1).

**Table 1.**
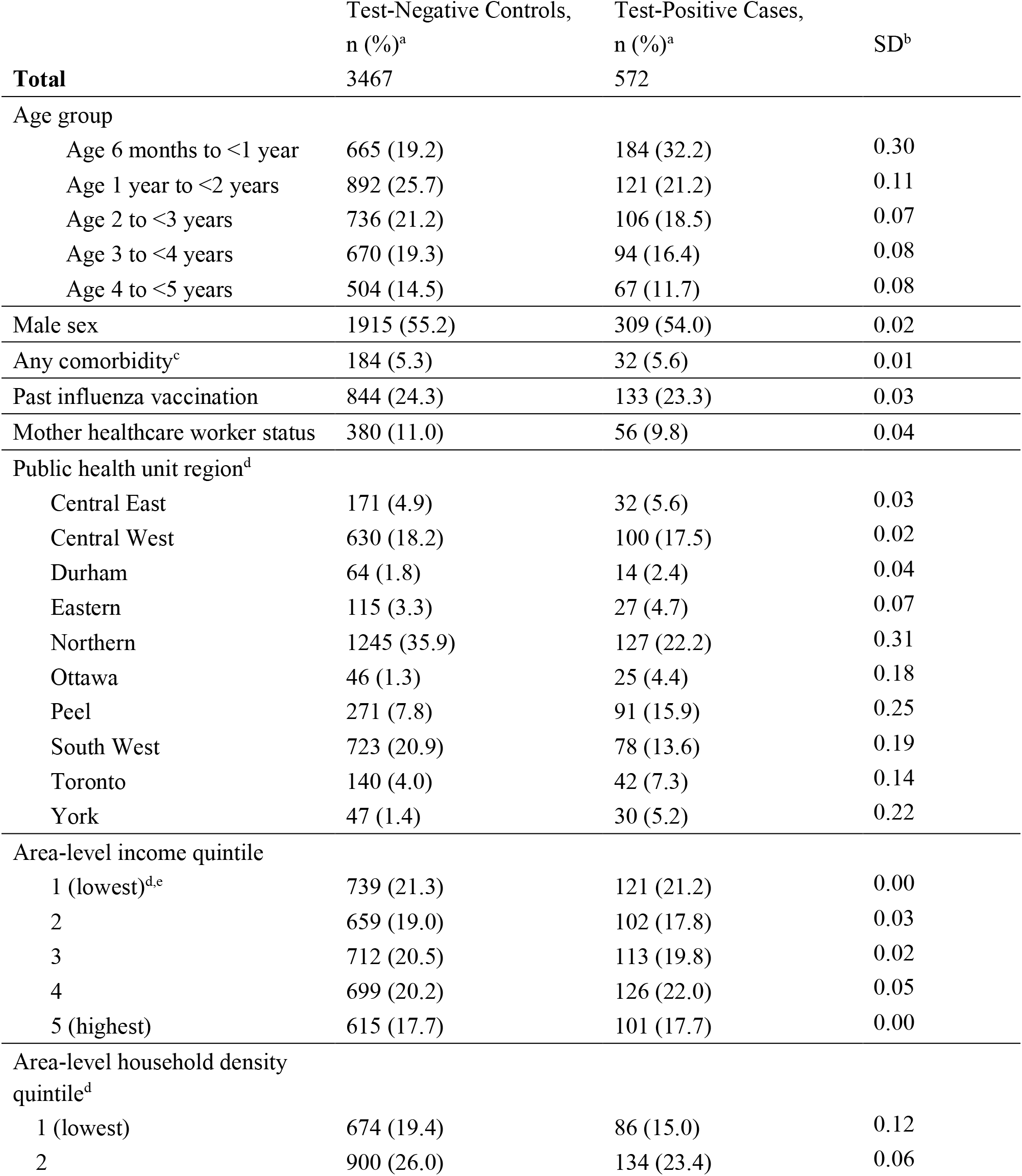
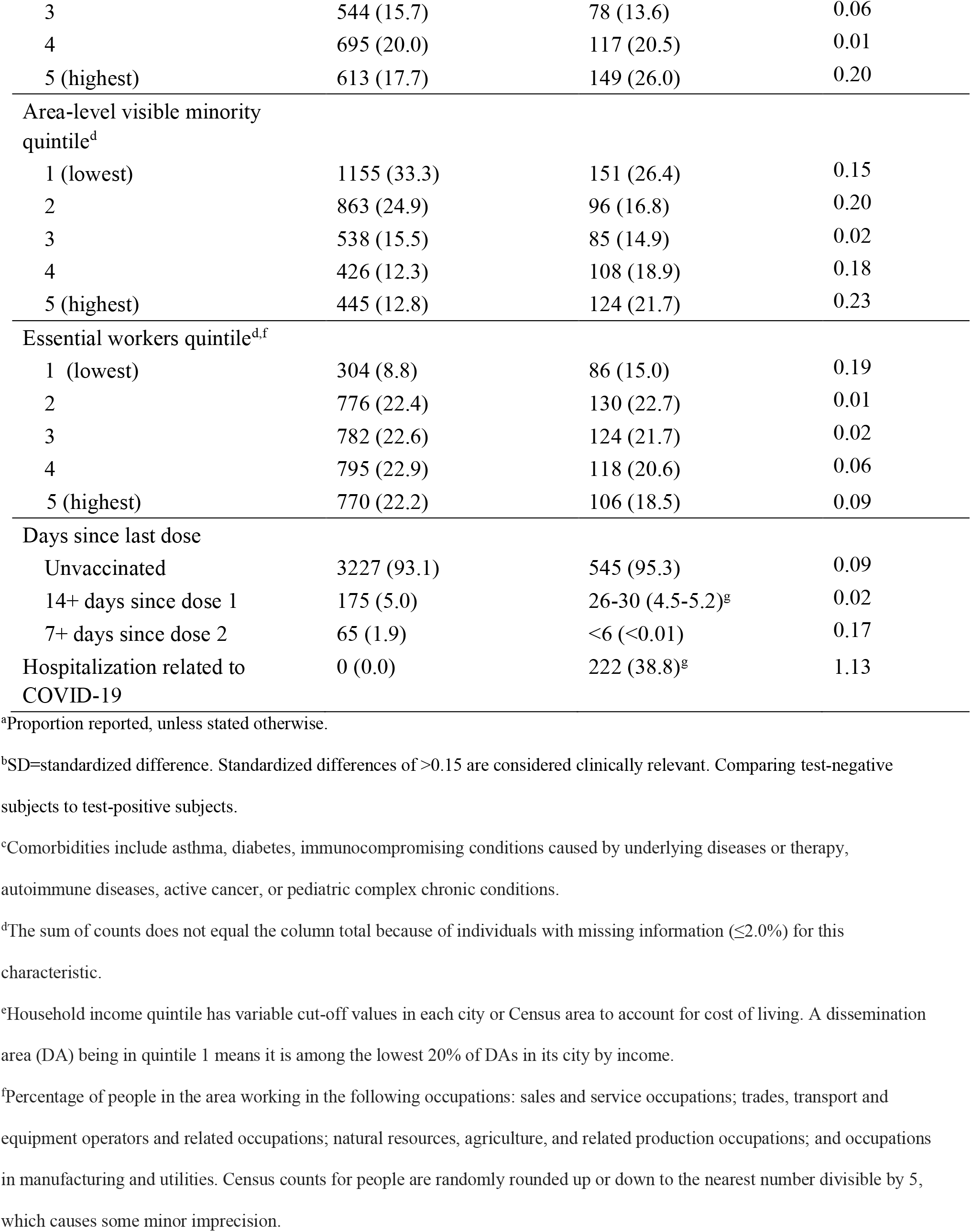
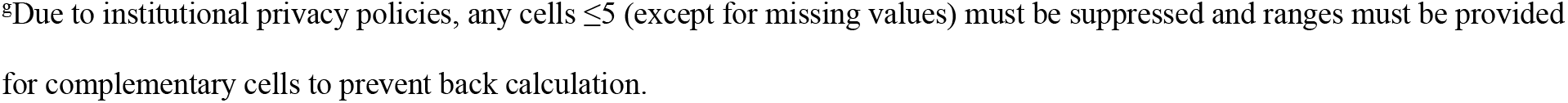
Descriptive characteristics of children aged 6 months to 5 years tested for SARS-CoV-2 with COVID-19-related symptoms between July 28, 2022, and December 31, 2022 comparing test-positive cases to test-negative controls (n=4,039)

|  | Test-Negative Controls,<br>n (%)<sup>a</sup> | Test-Positive Cases,<br>n (%)<sup>a</sup> | SD<sup>b</sup> |
| --- | --- | --- | --- |
| **Total** | 3467 | 572 |  |
| Age group |  |  |  |
| Age 6 months to <1 year | 665 (19.2) | 184 (32.2) | 0.30 |
| Age 1 year to <2 years | 892 (25.7) | 121 (21.2) | 0.11 |
| Age 2 to <3 years | 736 (21.2) | 106 (18.5) | 0.07 |
| Age 3 to <4 years | 670 (19.3) | 94 (16.4) | 0.08 |
| Age 4 to <5 years | 504 (14.5) | 67 (11.7) | 0.08 |
| Male sex | 1915 (55.2) | 309 (54.0) | 0.02 |
| Any comorbidity<sup>c</sup> | 184 (5.3) | 32 (5.6) | 0.01 |
| Past influenza vaccination | 844 (24.3) | 133 (23.3) | 0.03 |
| Mother healthcare worker status | 380 (11.0) | 56 (9.8) | 0.04 |
| Public health unit region<sup>d</sup> |  |  |  |
| Central East | 171 (4.9) | 32 (5.6) | 0.03 |
| Central West | 630 (18.2) | 100 (17.5) | 0.02 |
| Durham | 64 (1.8) | 14 (2.4) | 0.04 |
| Eastern | 115 (3.3) | 27 (4.7) | 0.07 |
| Northern | 1245 (35.9) | 127 (22.2) | 0.31 |
| Ottawa | 46 (1.3) | 25 (4.4) | 0.18 |
| Peel | 271 (7.8) | 91 (15.9) | 0.25 |
| South West | 723 (20.9) | 78 (13.6) | 0.19 |
| Toronto | 140 (4.0) | 42 (7.3) | 0.14 |
| York | 47 (1.4) | 30 (5.2) | 0.22 |
| Area-level income quintile |  |  |  |
| 1 (lowest)<sup>d,e</sup> | 739 (21.3) | 121 (21.2) | 0.00 |
| 2 | 659 (19.0) | 102 (17.8) | 0.03 |
| 3 | 712 (20.5) | 113 (19.8) | 0.02 |
| 4 | 699 (20.2) | 126 (22.0) | 0.05 |
| 5 (highest) | 615 (17.7) | 101 (17.7) | 0.00 |
| Area-level household density<br>quintile<sup>d</sup> |  |  |  |
| 1 (lowest) | 674 (19.4) | 86 (15.0) | 0.12 |
| 2 | 900 (26.0) | 134 (23.4) | 0.06 |
| 3 | 544 (15.7) | 78 (13.6) | 0.06 |
| 4 | 695 (20.0) | 117 (20.5) | 0.01 |
| 5 (highest) | 613 (17.7) | 149 (26.0) | 0.20 |
| Area-level visible minority quintile<sup>d</sup> |  |  |  |
| 1 (lowest) | 1155 (33.3) | 151 (26.4) | 0.15 |
| 2 | 863 (24.9) | 96 (16.8) | 0.20 |
| 3 | 538 (15.5) | 85 (14.9) | 0.02 |
| 4 | 426 (12.3) | 108 (18.9) | 0.18 |
| 5 (highest) | 445 (12.8) | 124 (21.7) | 0.23 |
| Essential workers quintile<sup>d,f</sup> |  |  |  |
| 1 (lowest) | 304 (8.8) | 86 (15.0) | 0.19 |
| 2 | 776 (22.4) | 130 (22.7) | 0.01 |
| 3 | 782 (22.6) | 124 (21.7) | 0.02 |
| 4 | 795 (22.9) | 118 (20.6) | 0.06 |
| 5 (highest) | 770 (22.2) | 106 (18.5) | 0.09 |
| Days since last dose |  |  |  |
| Unvaccinated | 3227 (93.1) | 545 (95.3) | 0.09 |
| 14+ days since dose 1 | 175 (5.0) | 26-30 (4.5-5.2)<sup>g</sup> | 0.02 |
| 7+ days since dose 2 | 65 (1.9) | <6 (<0.01) | 0.17 |
| Hospitalization related to COVID-19 | 0 (0.0) | 222 (38.8)<sup>g</sup> | 1.13 |

<sup>a</sup>Proportion reported, unless stated otherwise.
<sup>b</sup>SD=standardized difference. Standardized differences of >0.15 are considered clinically relevant. Comparing test-negative subjects to test-positive subjects.
<sup>c</sup>Comorbidities include asthma, diabetes, immunocompromising conditions caused by underlying diseases or therapy, autoimmune diseases, active cancer, or pediatric complex chronic conditions.
<sup>d</sup>The sum of counts does not equal the column total because of individuals with missing information ( $\leq 2.0\%$ ) for this characteristic.
<sup>e</sup>Household income quintile has variable cut-off values in each city or Census area to account for cost of living. A dissemination area (DA) being in quintile 1 means it is among the lowest 20% of DAs in its city by income.
<sup>f</sup>Percentage of people in the area working in the following occupations: sales and service occupations; trades, transport and equipment operators and related occupations; natural resources, agriculture, and related production occupations; and occupations in manufacturing and utilities. Census counts for people are randomly rounded up or down to the nearest number divisible by 5, which causes some minor imprecision.
§Due to institutional privacy policies, any cells $\leq 5$ (except for missing values) must be suppressed and ranges must be provided for complementary cells to prevent back calculation.

**Figure 1.**
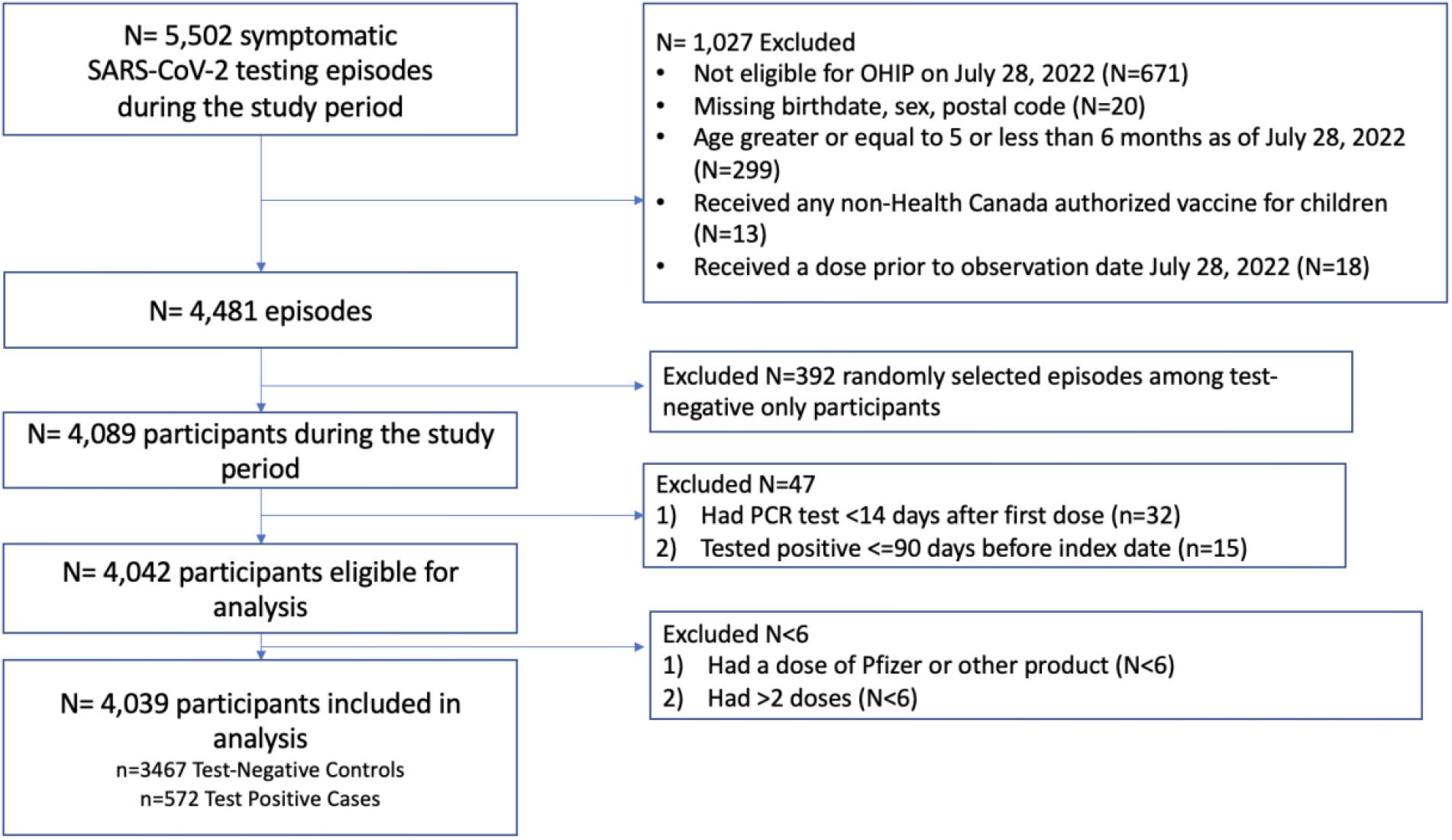
Flowchart of children aged 6 months to <5 years old tested for symptomatic SARS-CoV-2 infection during the study period from July 28 to December 31, 2022 OHIP=Ontario Health Insurance Plan PCR= Real-time polymerase chain reaction (RT-PCR)

Overall VE against symptomatic SARS-CoV-2 infection was 20% (95%CI: –27, 59%) ≥14 days following a first dose, and 90% (95%CI: 53, 99%) ≥7 days following a second dose (Figure 2).

**Figure 2.**
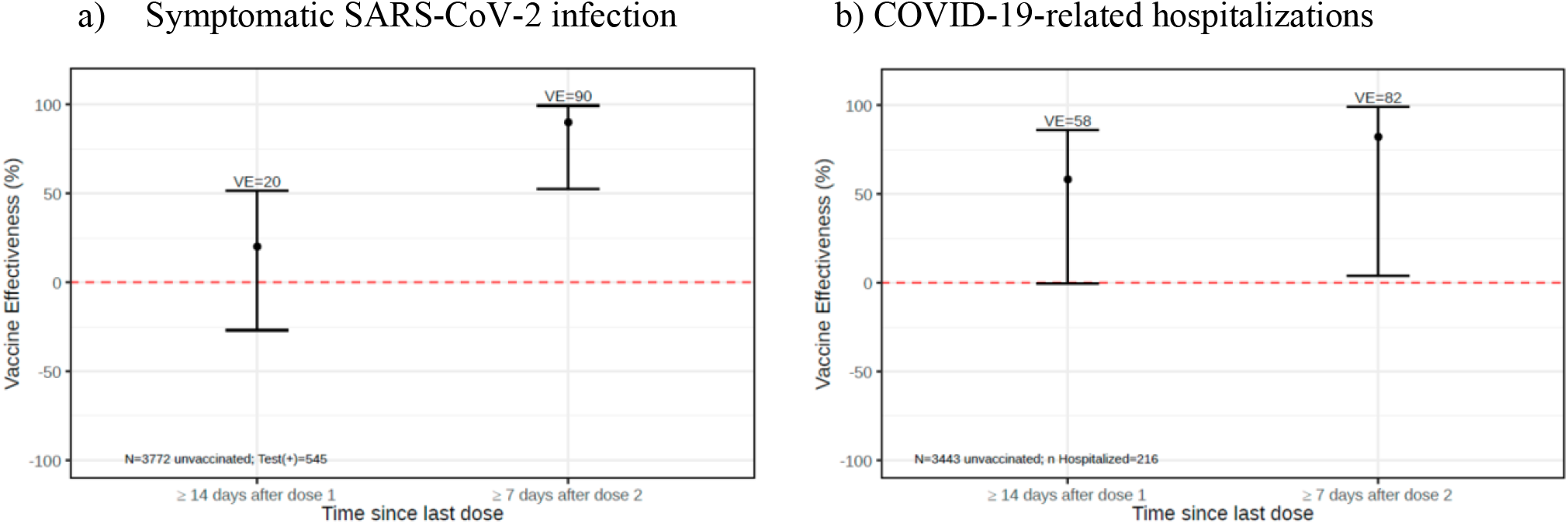
Vaccine effectiveness estimates^a^ in children aged 6 months to 5 years old against a) symptomatic SARS-CoV-2 infection and b) COVID-19-related hospitalizations a) Symptomatic SARS-CoV-2 infection b) COVID-19-related hospitalizations ^a^Confounders included in the model were week of test, age, sex, medical comorbidities, prior influenza vaccination, having a healthcare worker mother, positive SARS-CoV-2 RT-PCR test >= 90 days before the index date, household income quintile, household density quintile, essential workers quintile, public health unit, and number of physician visits (modelled as a non-linear continuous variable using restricted cubic splines).

VE for a single dose peaked at 68% (95%CI: 18, 90%) between 14 and 29 days after a first dose and declined thereafter (Supplementary Figure 1). We had insufficient sample size to assess VE at monthly intervals after a second dose.

We observed only a small number of COVID-19-related hospitalizations (n=222), with <10 among children who had received at least one dose of mRNA-1273. VE against COVID-19-related hospitalization was 58% (95%CI: 0, 86%) ≥14 days after a first dose, and 82% (95%CI: 4, 99%) ≥7 days after a second dose (Figure 2).

We did not detect heterogeneity of VE against symptomatic SARS-CoV-2 infection by age or previous documented SARS-CoV-2 infection (LRT P=0.56 and P=0.15, respectively).

## DISCUSSION

Our findings suggest that 2 doses of mRNA-1273 provide initial protection against symptomatic SARS-CoV-2 infection and COVID-19-related hospitalization in children aged 6 months to 5 years.

An earlier study of VE against symptomatic infection among uninsured children aged 3 to 5 years in the United States (n=37,010) found a VE of 40% (95%CI: 26, 52%) 2 weeks after a first dose, and a peak of 60% (95%CI: 49, 68%) 2 weeks after a second dose.^8^ VE against COVID-19-related hospitalization was not evaluated in that study. Compared to other studies in Ontario, Canada, VE against symptomatic infection was comparable to children aged 5-11 years after a first dose (20% vs 24% 14 to 29 days after a first dose), with higher VE after a second dose (90% vs 66% 7 to 29 days after a second dose).^4^ Although no deaths related to COVID-19 were recorded in our study, VE against either hospitalization or death associated with COVID-19 in children aged 5-11 years was comparable to our findings (82% vs 79% ≥ 7 days after a second dose).

Study limitations include potential residual confounding due to health-seeking behaviours.^9^ We were also limited to symptomatic infection using this design, which could be vulnerable to misclassification. We may have also lacked statistical power to detect effect heterogeneity by age or previous documented SARS-CoV-2 infection. Generalizability to immunocompromised children and children who were unable to access testing during the study period may be limited.

## Data Availability

The dataset from this study is held securely in coded form at ICES. While legal data sharing agreements between ICES and data providers (e.g., healthcare organizations and government) prohibit ICES from making the dataset publicly available, access may be granted to those who meet pre-specified criteria for confidential access, available at https://www.ices.on.ca/DAS.

## Code availability

The full dataset creation plan and underlying analytic code are available from the authors upon request, understanding that the computer programs may rely upon coding templates or macros that are unique to ICES and are therefore either inaccessible or may require modification.

## Acknowledgments

We would like to acknowledge Public Health Ontario for access to vaccination data from COVaxON case-level data from the Public Health Case and Contact Management Solution (CCM) and COVID-19 laboratory data, as well as assistance with data interpretation. We also thank the staff of Ontario’s public health units who are responsible for COVID-19 case and contact management and data collection within CCM. This document used data adapted from the Statistics Canada Postal CodeOM Conversion File, which is based on data licensed from Canada Post Corporation, and/or data adapted from the Ontario Ministry of Health Postal Code Conversion File, which contains data copied under license from ©Canada Post Corporation and Statistics Canada. Parts of this material are based on data and/or information compiled and provided by: MOH, CIHI, Statistics Canada, and IQVIA Solutions Canada Inc. The analyses, conclusions, opinions and statements expressed herein are solely those of the authors and do not reflect those of the funding or data sources; no endorsement is intended or should be inferred. Adapted from Statistics Canada, Canadian Census 2016. This does not constitute an endorsement by Statistics Canada of this product. We thank IQVIA Solutions Canada Inc. for use of their Drug Information File.

## Patient Consent Statement

ICES is a prescribed entity under Ontario’s Personal Health Information Protection Act (PHIPA). Section 45 of PHIPA authorizes ICES to collect personal health information, without consent, for the purpose of analysis or compiling statistical information with respect to the management of, evaluation or monitoring of, the allocation of resources to or planning for all or part of the health system. Projects that use data collected by ICES under section 45 of PHIPA, and use no other data, are exempt from REB review. The use of the data in this project is authorized under section 45 and approved by ICES’ Privacy and Legal Office.

## Dissemination to participants and related patient and public communities

Results of this study have been made available to the public on a preprint server and have been shared with the Ministry of Health and ICES. Following peer review publication, they will be further disseminated by ICES through news media and social media. It is not possible to send study results to participants because all personal identifying information has been removed from the dataset.

