## Supplementary Material for "mRNA-1273 vaccine effectiveness against symptomatic SARS-CoV-2 infection and COVID-19-related hospitalization in children aged 6 months to 5 years"

### Supplementary Appendix

This appendix has been provided by the authors to give readers additional information about their work.

**Supplementary Text:** Determination of symptom status at the time of SARS-CoV-2 testing

The Ontario Laboratories Information System (OLIS) includes open fields (specifically the Patient Note Clinical Information field or the observation code XON13543-4 [Patient symptoms]) which document whether individuals tested for SARS-CoV-2 were symptomatic at the time of testing. These character-based fields were initially separated by commas, slashes, semicolons, or ampersands, and the character text-strings were subsequently parsed and compiled.

Author JCK determined symptom classifications in OLIS as of January 9, 2023, that were likely be attributed to COVID-19. Terms that appeared infrequently (less than 25 times in total) were excluded. The values recorded in the symptoms fields were categorized as either "symptomatic" or "asymptomatic." We deliberately adopted a wide-ranging definition to capture all potentially relevant COVID-19 symptoms, including unusual symptoms and chronic conditions, based on our scientific and medical understanding of symptoms related to COVID-19.

Below is a list of terms identified as indicative of COVID-19 symptoms (classified as 'symptomatic'). In addition to these terms, we also included the term SYMPTOMATIC (or partial spellings thereof) or any mention of symptom onset.

", *, +VE RAPID, +VE RAPID TEST, +VE RAT, -, ., .ASY, 0, 0,  TASTE, 0.97, 0.98, 0.99, 01, 02, 03, 04, 05, 06, 07, 08, 09, 1, 10, 100, 100%, 101, 102, 11, 12, 13, 14, 15, 16, 17, 18, 19, 2, 20, 2020, 2020 OUT PT, 2020  OUT PT, 2020...OUT PT, 2021, 2021    OUT PT, 2021   IN PT, 2021   OUT PT, 2021 IN PT, 2021  OUT PT, 2021 - COUGH, 2021 - FEVER, 2021 - SORE THROAT, 2021-11-05, 2021-11-16, 2021-11-17, 2021-11-18, 2021-12-21, 2021-12-22, 2021-12-25, 2021-12-26, 2021-12-27, 2021-12-28, 2021-12-29, 2021., 2022, 2022   IN PT, 2022   OUT PT, 2022  IN PT, 2022  OUT PT, 21, 21 - COUGH, 21., 22, 23, 24, 25, 26, 27, 28, 29, 2ND SWAB, 3, 30, 31, 35, 35.0, 35.1, 35.2, 35.3, 35.4, 35.5, 35.6, 35.7, 35.8, 35.9, 36, 36.0, 36.1, 36.2, 36.3, 36.4, 36.5, 36.6, 36.7, 36.8, 36.9, 37, 37.0, 37.1, 37.2, 37.3, 37.4, 37.5, 37.6, 37.7, 37.8, 37.9, 38, 38.0, 38.1, 38.2, 38.3, 38.4, 38.5, 38.6, 38.7, 38.8, 38.9, 39, 39.0, 39.1, 39.2, 39.3, 39.4, 39.5, 39.6, 39.7, 3RD SWAB, 4, 40, 5, 6, 7, 8, 9, 97%, 98%, 99, 99%, A, A COLD, AB PAIN, ABD, ABD DISCOMFORT, ABD PAIN, ABD. PAIN, ABD.PAIN, ABDO CRAMPS, ABDO DISCOMFORT, ABDO PAIN, ABDO PAIN AND HEADACHE, ABDOMEN PAIN, ABDOMINAL, ABDOMINAL CRAMPING, ABDOMINAL CRAMPS, ABDOMINAL DISCOMFORT, ABDOMINAL PAIN, ABDOMINAL PAINS, ABDOMINAL UPSET, ABDOPAIN, ABNORMAL ULTRASOUND, AC VISITOR, ACHE, ACHES, ACHES AND CHILLS, ACHES AND PAIN, ACHES AND PAINS, ACHES CHILLS, ACHES FATIGUE, ACHES HEADACHE, ACHES RUNNY NOSE, ACHEY, ACHINESS, ACHING, ACHY, ACHY BODY, ACHY HEADACHE, ACHY JOINTS, ACHY MUSCLES, ACUTE STROKE, ADMISSION, ADMISSION ONLY, ADMISSION PROTOCOL, ADMISSION SWAB, ADMISSION TO HOSPICE, ADMISSION TO HOSPITAL, ADMISSON, ADMIT, ADMITTED, AFEBRILE, AFIB, ALLERGIES, ALLERGY SYMPTOMS, ALTERED LOC, ALTERED TASTE, AND HEADACHE, ANEMIA, ANOREXIA, ANOSMIA, ANXIETY, AP, APPENDICITIS, APPETITE, APRIL 22, APRIL 28, ARTHRALGIA, ARTHRITIS, AS, ASTHMA, ASX, ASY, ASYMPOTMATIC, ASYMPT., ASYMPTOMATIC, ASYMPTOMATIC - CLEARANCE, ASYMPTOMATIC - EXPOSURE, ASYMPTOMATIC - SURVEILLANCE, ASYPMTOMATIC, ASYPTOMATIC, AUG 30, AUTOPSY, AX CENTRE, AYMPTOMATIC, Anemia, Apr-22, Apr-28, B, BACK ACHE, BACK ACHES, BACK PAIN, BACKACHE, BACKPAIN, BAD HEADACHE, BARKING COUGH, BEING ADMITTED, BLEEDING, BLEEDING IN PREGNANCY, BLOATING, BLOODY STOOLS, BODY, BODY ACH, BODY ACHE, BODY ACHE AND HEADACHE, BODY ACHE HEADACHE, BODY ACHE., BODY ACHEA, BODY ACHES,

BODY ACHES 12, BODY ACHES AND CHILLS, BODY ACHES AND HEADACHE, BODY ACHES AND PAIN, BODY ACHES AND PAINS, BODY ACHES CHILLS, BODY ACHES CONGESTION, BODY ACHES FATIGUE, BODY ACHES HEADACHE, BODY ACHES HEADACHES, BODY ACHES RUNNY NOSE, BODY ACHES., BODY ACHES. 2022, BODY CHILLS, BODY MALAISE, BODY PAIN, BODY PAINS, BODY RASH, BODY WEAKNESS, BODYACH, BODYACHE, BODYACHES, BODYACHES CHILLS, BODYPAIN, BOSY ACHES, BRAIN FOG, BREATHING DIFFICULTY, BREATHING ISSUES, BRONCHITIS, BURNING CHEST, BURNING EYES, BURNING IN CHEST, Bleeding in pregnancy, Bloating, Bloody stools, C, C-CASE, CAMP, CANCER, CARDIAC, CARDIAC ARREST, CAREGIVER, CATARACT, CCU), CENTRAL CAREGIVER FOR LONG-TERM CARE, CEPHALGIA, CH, CHANGE IN SMELL, CHANGE IN TASTE, CHANGE IN TASTE AND SMELL, CHEST, CHEST AND NASAL CONGESTION, CHEST BURNING, CHEST COLD, CHEST CONG, CHEST CONGESTED, CHEST CONGESTION, CHEST DISCOMFORT, CHEST HEAVINESS, CHEST HEAVY, CHEST HURTS, CHEST INFECTION, CHEST IRRITATION, CHEST PAIN, CHEST PAINS, CHEST PRESSURE, CHEST SORE, CHEST SORENESS, CHEST TIGHNESS, CHEST TIGHT, CHEST TIGHTNESS, CHESTPAIN, CHF, CHI, CHIILS, CHIL, CHILL, CHILLA, CHILLLS, CHILLS, CHILLS ACHES, CHILLS AND BODY ACHE, CHILLS AND BODY ACHES, CHILLS AND FATIGUE, CHILLS AND HEADACHE, CHILLS AND RUNNY NOSE, CHILLS AND SWEATS, CHILLS BODY ACHE, CHILLS BODY ACHES, CHILLS CONGESTION, CHILLS DIARRHEA, CHILLS FATIGUE, CHILLS HEADACHE, CHILLS NAUSEA, CHILLS RUNNY NOSE, CHILLS SWEATS, CHILLS., CHILLS. 2021, CHILLS. 2022, CHILLS. HEADACHE, CHILS, CHRONIC COUGH, CLAMMY, CLEARANCE, CLEARING THROAT, CLOSE CONTACT, COGESTION, COGUH, COLD, COLD CHILLS, COLD FLASHES, COLD LIKE, COLD LIKE SYMPTOMS, COLD RUNNY NOSE, COLD SORE, COLD SWEAT, COLD SWEATS, COLD SX, COLD SYMPTOMS, COLD-LIKE SYMPTOMS, COLDS, CONFUSED, CONFUSION, CONG, CONGE, CONGEATION, CONGES, CONGESITON, CONGEST, CONGESTED, CONGESTED CHEST, CONGESTED COUGH, CONGESTED FATIGUE, CONGESTED HEADACHE, CONGESTED NOSE, CONGESTED RUNNY NOSE, CONGESTED SNEEZING, CONGESTED., CONGESTI, CONGESTIO, CONGESTION, CONGESTION 2021, CONGESTION ACHES, CONGESTION AND FATIGUE, CONGESTION AND HEADACHE, CONGESTION AND RUNNY NOSE, CONGESTION BODY ACHES, CONGESTION CHILLS, CONGESTION DIARRHEA, CONGESTION FATIGUE, CONGESTION FATIGUE HEADACHE, CONGESTION HA, CONGESTION HEADACHE, CONGESTION HEADACHES, CONGESTION NASAL, CONGESTION NAUSEA, CONGESTION RUNNY NOSE, CONGESTION RUNNY NOSE HEADACHE, CONGESTION SNEEZING, CONGESTION SOB, CONGESTION., CONGESTION. 2021, CONGESTION. 2022, CONGESTION. HEADACHE, CONGESTION. RUNNY NOSE, CONGESTIONS, CONGESTON, CONGETION, CONGSTION, CONJESTION, CONJUCTION, CONJUCTIVITIS, CONJUNCTION, CONJUNCTIVITIS, CONSTIPATION, CONTACT, COPD, CORE THROAT, CORYZA, COUGH, COUGH 12, COUGH 2021, COUGH AND SORE THROAT, COUGH CONGESTION, COUGH DRY, COUGH FEVER, COUGH ONSET 20, COUGH PRODUCTIVE, COUGH RUNNY NOSE, COUGH SOB, COUGH SORE THROAT, COUGH SORE THROAT RUNNY NOSE, COUGH., COUGH. 2021, COUGH. 2022, COUGHING, COVID POSITIVE, COVID RESWAB, COVID TESTING, CP, CRAMPING, CRAMPS, CROUP, D, DAIRRHEA, DATE :2021, DATE OF SYMPTOM ONSET: 2021, DATE: 2021, DATE:2021, DD, DEC, DEC APPETITE, DEC.19, DEC.20, DEC.21, DEC.22, DEC.23, DEC.24, DEC.25, DEC.26, DEC.27, DEC.APPETITE, DECEASED, DECREASE APPETITE, DECREASE APPETITE. 2021, DECREASE TASTE, DECREASED APPETITE, DECREASED APPETITE., DECREASED DRINKING, DECREASED LOC, DECREASED SMELL, DECREASED TASTE, DEHYDRATION, DELERIUM, DELIRIUM, DEMENTIA, DEPRESSION, DIA, DIAHERRA, DIAHHREA, DIAHREA, DIAHRREA, DIAPHORESIS, DIAPHORETIC, DIAR, DIAREHA, DIARHEA, DIARHHEA, DIARR, DIARREA, DIARREAH, DIARREHA, DIARREHEA, DIARRH, DIARRHE, DIARRHEA, DIARRHEA AND FATIGUE, DIARRHEA AND HEADACHE, DIARRHEA AND NAUSEA,

DIARRHEA AND VOMITING, DIARRHEA BLOODY, DIARRHEA CHILLS, DIARRHEA FATIGUE, DIARRHEA HEADACHE, DIARRHEA NAUSEA, DIARRHEA RUNNY NOSE, DIARRHEA VOMITING, DIARRHEA WATERY, DIARRHEA., DIARRHEA. 2021, DIARRHOEA, DIARROHEA, DIFF BREATHING, DIFF SWALLOWING, DIFFICULT BREATHING, DIFFICULT SWALLOWING, DIFFICULTY, DIFFICULTY BREATHING, DIFFICULTY IN BREATHING, DIFFICULTY SWALLOWING, DIGESTIVE ISSUES, DIRECT CONTACT, DIRRHEA, DISCHARGE, DISCOMFORT, DIZINESS, DIZZINES, DIZZINESS, DIZZY, DIZZY HEADACHE, DIZZY NAUSEA, DIZZYNESS, DKA, DOB, DR APRIASZ, DR RUTKA, DROWSINESS, DROWSY, DRY, DRY COUGH, DRY EYES, DRY MOUTH, DRY NOSE, DRY THROAT, DYSPEPSIA, DYSPHAGIA, DYSPNEA, DYSURIA, Dysuria, EAR, EAR ACHE, EAR ACHES, EAR CONGESTION, EAR INFECTION, EAR PAIN, EAR PRESSURE, EARACHE, EARACHES, EARPAIN, EARS, EARS HURT, EARS PLUGGED, ED, ED PATIENT, ED PT, ELEVATED LIVER ENZYMES OR LIVER FUNCTION TESTS, EMERGE DEPT, EMERGENCY DEPT, EMESIS, EMESIS DIARRHEA, EMESIS X 1, EMESIS X 2, EMESIS X1, EMP HEALTH, EMPLOYEE, EMPLOYEE HEALTH, EMPLOYEE/ STAFF MEMBER, EMS, EMS ASYMP, EMS OUTREACH, EMS SYMP, ENCEPHALITIS, ENDO, EPIGASTRIC PAIN, ER, ER - TO BE HOSPITALIZED, ER ADMIT, ER PATIENT, ER PATIENT ADMITTED, ER PT, ER PT ADMITTED, ERROR, ESSENTIAL CAREGIVER, ESSENTIAL CAREGIVER FOR LONG-TERM CARE, EXHAUSTED, EXHAUSTION, EXPOSED, EXPOSED AND PREGNANT, EXPOSURE, EXPOSURE TO MICE, EXTREME FATIGUE, EXTREME TIREDNESS, EYE, EYE DISCHARGE, EYE INFECTION, EYE IRRITATION, EYE PAIN, EYE REDNESS, EYES, EYES BURNING, EYES HURT, Encephalitis, F, FA, FAILURE TO COPE, FAINT, FAITGUE, FAITUGE, FALL, FALLS, FAT, FATGIUE, FATGUE, FATI, FATIG, FATIGE, FATIGU, FATIGUE, FATIGUE 12, FATIGUE 2021, FATIGUE ACHES, FATIGUE AND BODY ACHES, FATIGUE AND CONGESTION, FATIGUE AND HEADACHE, FATIGUE AND NAUSEA, FATIGUE AND RUNNY NOSE, FATIGUE BODY ACHE, FATIGUE BODY ACHES, FATIGUE CHILLS, FATIGUE CONGESTED, FATIGUE CONGESTION, FATIGUE DIARRHEA, FATIGUE HEADACHE, FATIGUE MALAISE, FATIGUE MUSCLE ACHES, FATIGUE MYALGIA, FATIGUE NASAL CONGESTION, FATIGUE NAUSEA, FATIGUE RUNNY NOSE, FATIGUE SOB, FATIGUE., FATIGUE. 2021, FATIGUE. 2022, FATIGUE. HEADACHE, FATIGUED, FATIGUES, FATIQUE, FATTY LIVER, FATUGUE, FATUIGE, FEB 8 OR, FEBRILE, FEELING COLD, FEELING FEVERISH, FEELING HOT, FEELING TIRED, FEELING UNWELL, FEELING WARM, FEELING WEAK, FEELS FEVERISH, FEELS WARM, FELT FEVERISH, FELT WARM, FERTILITY, FEVER, FEVER 37, FEVER 37.8, FEVER 37.9, FEVER 38, FEVER 38.1, FEVER 38.2, FEVER 38.5, FEVER AT HOME, FEVER COUGH, FEVER COUGH SORE THROAT, FEVER LAST NIGHT, FEVER NYD, FEVER RESOLVED, FEVER TODAY, FEVER., FEVER. 2021, FEVERISH, FEVERS, FEVR, FLANK PAIN, FLEM, FLU, FLU LIKE, FLU LIKE SYMPTOMS, FLU SYMPTOMS, FLU-LIKE SYMPTOMS, FLUSHED, FOGGY, FOGGY HEAD, FOR ADMISSION, FOR DISCHARGE, FOR LTC VISIT, FOR OR, FOR PLACEMENT, FOR PRE OP, FOR PRECAUTION, FOR PROCEDURE, FOR SAFETY, FOR SCHOOL, FOR SURGERY, FOR TRANSFER, FOR WORK, FOR WORK., FULL VAC, FULL VACC, FULL VACC., FULLY VACCINATED., FUSSY, GASTRITIS, GASTRO, GASTRO ISSUES, GASTRO SYMPTOMS, GASTROENTERITIS, GASTROINTESTINAL, GEN UNWELL, GEN WEAKNESS, GENERAL FATIGUE, GENERAL MALAISE, GENERAL UNWELL, GENERAL WEAKNESS, GENERALIZED WEAKNESS, GENERALLY UNWELL, GERD, GI, GI BLEED, GI BLEEDING, GI ISSUE, GI ISSUES, GI SYMPTOMS, GI UPSET, GI bleeding, GOING TO OR, GREEN PHLEGM, GREEN SPUTUM, H, H.VOICE, HA, HA ACHES, HA RN, HA., HADACHE, HALLUCINATIONS, HARD TO BREATH, HARD TO BREATHE, HARD TO SWALLOW, HCV RE-EXPOSURE, HCW, HCW-SURVEILLANCE, HEA, HEAACHE, HEAADCHE, HEACHACHE, HEACHE, HEAD, HEAD ACHE, HEAD ACHES, HEAD AND BODY ACHE, HEAD COLD, HEAD CONGESTION, HEAD PRESSURE, HEADA, HEADAACHE, HEADAC, HEADACE, HEADACEH, HEADACH, HEADACHE, HEADACHE  9, HEADACHE (ONLY), HEADACHE 12, HEADACHE 2021,

HEADACHE ACHES, HEADACHE ACHY, HEADACHE AND BODY ACHE, HEADACHE AND BODY ACHES, HEADACHE AND BODY PAIN, HEADACHE AND CHILLS, HEADACHE AND CONGESTION, HEADACHE AND DIARRHEA, HEADACHE AND FATIGUE, HEADACHE AND NASAL CONGESTION, HEADACHE AND NAUSEA, HEADACHE AND RUNNY NOSE, HEADACHE AND STUFFY NOSE, HEADACHE AND VOMITING, HEADACHE BODY ACHE, HEADACHE BODY ACHES, HEADACHE BODY PAIN, HEADACHE BODYACHE, HEADACHE CHEST PAIN, HEADACHE CHILLS, HEADACHE CONGESTED, HEADACHE CONGESTION, HEADACHE DIARRHEA, HEADACHE DIZZINESS, HEADACHE DIZZY, HEADACHE FATIGUE, HEADACHE FATIGUE NAUSEA, HEADACHE FATIGUED, HEADACHE LOSS OF TASTE, HEADACHE MALAISE, HEADACHE MUSCLE ACHE, HEADACHE MUSCLE ACHES, HEADACHE MUSCLE PAIN, HEADACHE MYALGIA, HEADACHE NASAL CONGESTION, HEADACHE NAUSEA, HEADACHE NAUSEA DIARRHEA, HEADACHE NAUSEA FATIGUE, HEADACHE NAUSEA VOMITING, HEADACHE RHINORRHEA, HEADACHE RUNNY NOSE, HEADACHE RUNNY NOSE FATIGUE, HEADACHE SINUS, HEADACHE SINUS CONGESTION, HEADACHE SNEEZING, HEADACHE SOB, HEADACHE STOMACH ACHE, HEADACHE STUFFY NOSE, HEADACHE TIRED, HEADACHE UPSET STOMACH, HEADACHE VOMITING, HEADACHE VSS, HEADACHE WEAKNESS, HEADACHE., HEADACHE. 2021, HEADACHE. 2022, HEADACHE. CHILLS, HEADACHE. FATIGUE, HEADACHE. NAUSEA, HEADACHE. RUNNY NOSE, HEADACHE.VSS, HEADACHE/STIFF NECK, HEADACHES, HEADACHES BODY ACHES, HEADACHES FATIGUE, HEADACHES RUNNY NOSE, HEADACHES., HEADAHCE, HEADAHE, HEADCAHE, HEADCHE, HEADCOLD, HEALTH CARE WORKER, HEALTHCARE WORKER, HEART FAILURE, HEART PALPITATIONS, HEARTBURN, HEAVINESS IN CHEST, HEAVY BREATHING, HEAVY CHEST, HEAVY HEAD, HEDACHE, HEMATURIA, HEMOPTYSIS, HIA, HIGH RISK, HIGH RISK CONTACT, HIVES, HOARSE, HOARSE THROAT, HOARSE VOI, HOARSE VOICE, HOARSENESS, HOARSENESS OF VOICE, HOMELESS, HORSE VOICE, HOT, HOT AND COLD, HOT AND COLD FLASHES, HOT FLASHES, HR-70, HURTS TO SWALLOW, HYPOXIA, Heart failure, Hematuria, IMMUNOCOMPROMISED, IN LABOUR, IN-PT, INCREASED SOB, INCREASED WOB, INDIGESTION, INDIRECT CONTACT, INFASS, INFASS OUTPATIENT, INFERTILITY, INFORMATION, INPATIENT, INPATIENT (HOSPITALIZED), INPATIENT (ICU, INPATIENT (ICU), INPT, INSOMNIA, INTUBATED, INUIT, IRRITABLE, IRRITATED EYES, IRRITATED THROAT, ITCHY, ITCHY EARS, ITCHY EYE, ITCHY EYES, ITCHY NOSE, ITCHY THROAT, IVF (IN VITRO FERTILIZATION), JAIL, JAN, JAUNDICE, JAW PAIN, JOINT ACHES, JOINT PAIN, JOINT PAINS, KCA, LABOUR, LABOURED BREATHING, LACK OF APPETITE, LACK OF ENERGY, LACK OF TASTE, LACK OF TASTE AND SMELL, LARYNGITIS, LATHARGIC, LBM, LEFT EAR PAIN, LEG PAIN, LETHARGIC, LETHARGY, LETHARY, LIGHT HEADACHE, LIGHT HEADED, LIGHT HEADEDNESS, LIGHT-HEADED, LIGHTHEADED, LIGHTHEADEDNESS, LOA, LONG-TERM CARE VISIT, LOOSE BM, LOOSE BOWEL, LOOSE BOWEL MOVEMENT, LOOSE BOWELS, LOOSE STOOL, LOOSE STOOLS, LOS, LOSE BOWEL MOVEMENT, LOSING VOICE, LOSS, LOSS APPETITE, LOSS OF APETITE, LOSS OF APPETITE, LOSS OF APPITITE, LOSS OF SENSE OF SMELL, LOSS OF SENSE OF TASTE, LOSS OF SENSE OF TASTE AND SMELL, LOSS OF SMELL, LOSS OF SMELL AND TASTE, LOSS OF SMELL OR TASTE, LOSS OF TASTE, LOSS OF TASTE AND SMELL, LOSS OF TASTE AND SMELL., LOSS OF TASTE OR SMELL, LOSS OF TASTE SMELL, LOSS OF VOICE, LOSS SMELL, LOSS SMELL AND TASTE, LOSS TASTE, LOSS TASTE AND SMELL, LOSS VOICE, LOST OF APPETITE, LOST OF SMELL, LOST OF TASTE, LOST OF TASTE AND SMELL, LOST TASTE, LOST TASTE AND SMELL, LOST VOICE, LOT, LOW APPEPTITE, LOW APPETITE, LOW BACK PAIN, LOW ENERGY, LOW FEVER, LOW GRADE FEVER, LOW RISK, LOWER BACK PAIN, LTC, LTC CLEARANCE, LTC EMPLOYEE, LTC HOME VISIT, LTC PLACEMENT, LTC RESIDENT, LTC SCREEN, LTC SCREENING, LTC VISIT, LTC VISITOR, LTC VISITS, LTC VIST, LTC WORKER, LTCWORKER, LUNG PAIN, LW INFASS, LW INFASS, LW INFASS (ASYMP), LW INFASS (SYMP), LW INFASS

ASYMP, LW INFASS OUTPATIENT, LW INFASS SYMP, LW INFASS SYMPT, LW INFASS SYMPT., LW INFASS: COUGH, M, MACULOPAPULAR RASH, MALAISE, MALASIE, MAY 6, MENINGITIS, METALLIC TASTE, MICE DROPPINGS, MIGRAINE, MIGRAINES, MIGRANE, MILD CHEST PAIN, MILD CONGESTION, MILD COUGH, MILD FEVER, MILD HEADACHE, MILD NASAL CONGESTION, MILD RUNNY NOSE, MILD SOB, MILD SORE THROAT, MUCOUS, MUCUS, MUSCLE, MUSCLE ACHE, MUSCLE ACHES, MUSCLE ACHES AND PAINS, MUSCLE ACHES FATIGUE, MUSCLE ACHES., MUSCLE AND JOINT PAIN, MUSCLE FATIGUE, MUSCLE PAIN, MUSCLE PAINS, MUSCLE SORE, MUSCLE SORENESS, MUSCLE WEAKNESS, MUSCLEACHE, MUSCLEACHES, MUSCLES ACHES, MYALGIA, MYALGIA HEADACHE, MYALGIA., MYALGIA. 2021, MYALGIAS, MYALGIAS-JOINT PAIN, May-06, Meningitis, N, N+V, NA, NARCOTIC ADDICTION, NAS, NAS.CONG, NASAL, NASAL AND CHEST CONGESTION, NASAL CON, NASAL CONG, NASAL CONG., NASAL CONGES, NASAL CONGESITON, NASAL CONGEST, NASAL CONGESTED, NASAL CONGESTI, NASAL CONGESTIO, NASAL CONGESTION, NASAL CONGESTION AND FATIGUE, NASAL CONGESTION AND HEADACHE, NASAL CONGESTION AND RUNNY NOSE, NASAL CONGESTION FATIGUE, NASAL CONGESTION HEADACHE, NASAL CONGESTION RUNNY NOSE, NASAL CONGESTION SNEEZING, NASAL CONGESTION., NASAL CONGESTION. 2021, NASAL CONGESTION. 2022, NASAL CONGESTIONS, NASAL CONGSTION, NASAL CONJESTION, NASAL DISCHARGE, NASAL DRAINAGE, NASAL DRIP, NASAL SYMPTOMS, NASALCONGESTION, NASEAU, NASEL CONGESTION, NASIA, NASUEA, NAU, NAUAEA, NAUS, NAUSA, NAUSE, NAUSEA, NAUSEA AND DIARRHEA, NAUSEA AND FATIGUE, NAUSEA AND HEADACHE, NAUSEA AND VOMITING, NAUSEA AND VOMITTING, NAUSEA CHILLS, NAUSEA DIARRHEA, NAUSEA FATIGUE, NAUSEA HEADACHE, NAUSEA RUNNY NOSE, NAUSEA VOMIT, NAUSEA VOMITING, NAUSEA VOMITING DIARRHEA, NAUSEA VOMITING HEADACHE, NAUSEA VOMITTING, NAUSEA., NAUSEAS, NAUSEATED, NAUSEAU, NAUSEOUS, NAUSIA, NECK PAIN, NEW ADMISSION, NEW ADMIT, NEW COUGH, NEW SMELL, NG, NIGHT SWEATS, NIL, NO, NO APPETITE, NO COUGH, NO ENERGY, NO FEVER, NO PNEUMONIA, NO SENSE OF SMELL, NO SENSE OF TASTE, NO SMELL, NO SMELL AND TASTE, NO SMELL OR TASTE, NO SOB, NO SORE THROAT, NO SYMPTOMS, NO SYMPTOMS NOTED, NO TASTE, NO TASTE AND SMELL, NO TASTE NO SMELL, NO TASTE OR SMELL, NO VOICE, NON-SPECIFIC SYMPTOM(S) - SURVEILLANCE, NONE, NONE LISTED, NORRHEA, NOSE, NOSE BLEED, NOSE CONGESTION, NOT APPLICABLE, NOT EATING, NOT FEELING WELL, NOT FOR TRAVEL, NOT GIVEN, NOT IMMUNIZED OR INCOMPLETE, NOT PROVIDED, NOT SPECIFIED, NOT VACCINATED, NOV, NS, NSTEMI, NURSING HOME, NURSING STUDENT, NV, NVD, NYD, N\T\V, O, O COVID, O2-97, O2-97%, O2-98, O2-98%, O2-99, O2-99%, OCCASIONAL COUGH, ONSET 20, ONSET 2020, ONSET UNKNOWN, ONSET YYYY-MM-DD, ONSET: 2021, OPP, OPP OFFICER, OR, OR TODAY, OTALGIA, OTHER, OTHER (SPECIFY), OUT PT, OUT-PT, OUTBREAK, OUTBREAK INVESTIGATION, OUTPATIENT, OUTPT, OVERDOSE, PAIN, PAIN IN CHEST, PAINS, PALPITATIONS, PANCREATITIS, PARENTS 2X VACC, PART VACC, PATIENT HAVING SURGERY, PEACE OF MIND, PFT, PHELGM, PHLEGM, PHLEGM IN THROAT, PHLEGMY, PHLEM, PINK EYE, PINK EYES, PINKEYE, PLACEMENT, PLUGGED EARS, PND, PNEUMONIA, PNEUMONIA (UNKNOWN), POOR APPETITE, POSITIVE ON RAPID TEST, POSITIVE RAPID, POSITIVE RAPID TEST, POSITIVE RAT, POSSIBLE EXPOSURE, POST NASAL, POST NASAL DRIP, POST PARTUM, PRE CARDIAC CATH, PRE CATH, PRE OP, PRE OP 2020, PRE OP SEPT 16, PRE OP SEPT 30, PRE PROCEDURE, PRE SURGICAL, PRE- OP, PRE-OP, PRE-OP 20, PRE-OP 2021, PRE-PROCEDURE, PRE-SURGICAL, PRECAUTION, PREGNANT, PREGNANT (LABOUR), PREOP, PRESSURE, PRESSURE IN CHEST, PRESSURE IN HEAD, PRESURGICAL, PREVIOUS POSITIVE, PREVIOUS RESULT INDETERMINATE, PROD COUGH, PRODUCTIVE COUGH, PUFFY EYES, Palpitations, R, R NOSE, R.NOSE, RAPID POSITIVE, RAPID TEST POSITIVE, RASH, RASH - NOT SPECIFIED, RASHES, RASPY THROAT, RASPY VOICE, RECEIVED ALL

DOSES > 14 DAYS AGO, RECENT FEVER, RECENT TRAVEL, RED EYE, RED EYES, REFUGEE, REGULAR TESTING, REMOTE COMMUNITY, RENAL CLINIC PATIENT, RENAL FAILURE, RENAL FAILURE/RENAL CLINIC PATIENT, REQUIRED BY PUBLIC HEALTH, REQUIRED FOR SURGERY, REQUIRED FOR WORK, RESOLVED, RESPIRATORY SYMPTOMS, RESTLESS, RESWAB, RETEST, RETRACTIONS, RETURN TO WORK, RETURNING, RHI, RHINITIS, RHINNORHEA, RHINNORRHEA, RHINO, RHINORHEA, RHINORR, RHINORREA, RHINORRHE, RHINORRHEA, RHINORRHEA CONGESTED, RHINORRHEA HEADACHE, RHINORRHEA-NASAL CONGESTION, RHINORRHEA., RHINORRHEA. 2021, RHIONRHEA, RIGHT EAR PAIN, RINGING IN EARS, RN, RN HA, RNNY NOSE, ROUTINE, RULE OUT COVID 19, RUN NOSE, RUNN YNOSE, RUNNING NOSE, RUNNING NOSE., RUNNING NOSE.VSS, RUNNING NOSR, RUNNNY NOSE, RUNNT NOSE, RUNNU NOSE, RUNNY, RUNNY  NOSE, RUNNY AND STUFFY NOSE, RUNNY CONGESTED NOSE, RUNNY EYES, RUNNY N, RUNNY NISE, RUNNY NO, RUNNY NOAE, RUNNY NOE, RUNNY NOISE, RUNNY NOS, RUNNY NOSE, RUNNY NOSE  9, RUNNY NOSE 11, RUNNY NOSE 12, RUNNY NOSE 2021, RUNNY NOSE ACHES, RUNNY NOSE AND BODY ACHE, RUNNY NOSE AND BODY ACHES, RUNNY NOSE AND CHILLS, RUNNY NOSE AND CONGESTED, RUNNY NOSE AND CONGESTION, RUNNY NOSE AND DIARRHEA, RUNNY NOSE AND EYES, RUNNY NOSE AND FATIGUE, RUNNY NOSE AND HEAD ACHE, RUNNY NOSE AND HEADACHE, RUNNY NOSE AND NASAL CONGESTION, RUNNY NOSE AND SNEEZING, RUNNY NOSE BODY ACHE, RUNNY NOSE BODY ACHES, RUNNY NOSE CHEST CONGESTION, RUNNY NOSE CHILLS, RUNNY NOSE CONGESTED, RUNNY NOSE CONGESTION, RUNNY NOSE CONGESTION HEADACHE, RUNNY NOSE COUGH, RUNNY NOSE DIARRHEA, RUNNY NOSE FATIGUE, RUNNY NOSE FATIGUE HEADACHE, RUNNY NOSE HEAD ACHE, RUNNY NOSE HEADACHE, RUNNY NOSE HEADACHE FATIGUE, RUNNY NOSE HEADACHES, RUNNY NOSE HOARSE VOICE, RUNNY NOSE LOSS OF TASTE, RUNNY NOSE MUSCLE ACHES, RUNNY NOSE NASAL CONGESTI, RUNNY NOSE NASAL CONGESTION, RUNNY NOSE NAUSEA, RUNNY NOSE ONSET 20, RUNNY NOSE OR NASAL CONGESTION, RUNNY NOSE OR SNEEZING, RUNNY NOSE SNEEZING, RUNNY NOSE SOB, RUNNY NOSE SORE THROAT, RUNNY NOSE STUFFY NOSE, RUNNY NOSE TIRED, RUNNY NOSE VOMITING, RUNNY NOSE VS NA, RUNNY NOSE WATERY EYES, RUNNY NOSE., RUNNY NOSE. 2021, RUNNY NOSE. 2022, RUNNY NOSE. CONGESTION, RUNNY NOSE. FATIGUE, RUNNY NOSE. HEADACHE, RUNNY NOSE. SNEEZING, RUNNY NOSE. T-36.0, RUNNY NOSES, RUNNY NOSR, RUNNY NOSW, RUNNY NOZE, RUNNY NSOE, RUNNY OSE, RUNNY ROSE, RUNNY STUFFY NOSE, RUNNYNOSE, RUNY NOSE, RUUNY NOSE, RYNNY NOSE, S, SCFHT, SCHIZOPHRENIA, SCHOOL, SCHOOL REQUIREMENT, SCRATCH THROAT, SCRATCHY, SCRATCHY THROAT, SCREENING, SEASONAL ALLERGIES, SEIZURE, SELF ASMT, SELF ASSESSMENT, SELF ASSMT, SEPSIS, SEVERE HEADACHE, SHAKES, SHAKING, SHAKY, SHELTER, SHIVERING, SHIVERS, SHORT BREATH, SHORT OF BREATH, SHORTNESS OF BREATH, SHORTNESS OF BREATH., SHORTNESS OF BREATHE, SHOULDER PAIN, SINUS, SINUS COLD, SINUS CONGESTED, SINUS CONGESTION, SINUS CONGESTION HEADACHE, SINUS HEADACHE, SINUS INFECTION, SINUS ISSUES, SINUS PAIN, SINUS PRESSURE, SINUS SYMPTOMS, SINUSES, SINUSITIS, SLEEPY, SLIGHT COUGH, SLIGHT FEVER, SLIGHT HEADACHE, SLIGHT RUNNY NOSE, SLIGHT SORE THROAT, SLUGGISH, SMELL, SMELL TASTE DISORDER, SNEEZ, SNEEZE, SNEEZING, SNEEZING AND RUNNY NOSE, SNEEZING CONGESTION, SNEEZING HEADACHE, SNEEZING RUNNY NOSE, SNEEZING., SNEEZY, SNEZZING, SNIFFING, SNIFFLE, SNIFFLES, SNIFFLING, SOB, SOB CONGESTION, SOB COUGH, SOB FATIGUE, SOB HEADACHE, SOB ON EXERTION, SOB RUNNY NOSE, SOB TODAY, SOB UPON EXERTION, SOB., SOBE, SOBOE, SOR ETHROAT, SOR THROAT, SORE, SORE BACK, SORE BODY, SORE CHEST, SORE EAR, SORE EARS, SORE EYE, SORE EYES, SORE JOINTS, SORE LEGS, SORE MUSCLE, SORE MUSCLES, SORE NECK, SORE STOMACH, SORE THOAT, SORE THORAT, SORE THRAOT, SORE THROAT, SORE THROAT 12, SORE THROAT 2021, SORE THROAT COUGH, SORE THROAT ONSET 20, SORE

THROAT RUNNY NOSE, SORE THROAT., SORE THROAT. 2021, SORE THROAT. 2022, SORE THT, SORE TUMMY, SORENESS, SORETHROAT, SORETHT, SPUTUM, STAFF, STAFF MEMBER, STEMI, STHROAT, STI, STIFF NECK, STIFFNESS, STOMACH, STOMACH ACHE, STOMACH ACHES, STOMACH BUG, STOMACH CRAMPS, STOMACH DISCOMFORT, STOMACH FLU, STOMACH HURTS, STOMACH ISSUES, STOMACH PAIN, STOMACH PAINS, STOMACH UPSET, STOMACHACHE, STOMACHE, STOMACHE ACHE, STREP, STREP THROAT, STROKE, STUFF NOSE, STUFF Y NOSE, STUFFED NOSE, STUFFED UP, STUFFED UP NOSE, STUFFINESS, STUFFING NOSE, STUFFY, STUFFY  NOSE, STUFFY AND RUNNY NOSE, STUFFY HEAD, STUFFY NOSE, STUFFY NOSE AND HEADACHE, STUFFY NOSE HEADACHE, STUFFY NOSE RUNNY NOSE, STUFFY NOSE SNEEZING, STUFFY NOSE., STUFFY NOSE. 2021, STUFFY RUNNY NOSE, STUFFYNOSE, STUFY NOSE, SUFFY NOSE, SUICIDAL, SURGERY, SURVEILLANCE, SURVIELLANCE, SWAB PRIOR TO CHEMO, SWAB PRIOR TO RADIATION, SWAB PRIOR TO STARTING CHEMO, SWAB PRIOR TO STARTING RADIATION, SWALLOWING, SWEAT, SWEATING, SWEATS, SWEATS AND CHILLS, SWEATY, SWELLING, SWOLLEN EYES, SWOLLEN GLAND, SWOLLEN GLANDS, SWOLLEN LYMPH NODES, SWOLLEN THROAT, SWOLLEN TONSILS, SX, SY, SYM, SYMP, SYMP-COUGH, SYMPOTMATIC, SYMPT, SYMPTOMATIC, SYMPTOMS, SYMPTOMS UNKNOWN, SYNCOPE, Sepsis, Sneezing, Stroke, T, T 36.0, T 36.1, T 36.2, T 36.3, T 36.4, T 36.5, T 36.6, T 36.7, T 36.8, T N, T+0, T+1, T+2, T+2 ENDO, T+3, T+4, T+5, T-1, T-35.2, T-35.8, T-36.0, T-36.1, T-36.2, T-36.3, T-36.4, T-36.5, T-36.6, T-N, T35.2, T35.6, T35.8, T35.9, T36, T36.1, T36.2, T36.3, T36.4, T36.5, T36.6, T36.7, T36.8, T36.9, TACHYCARDIA, TACHYPNEA, TASTE, TASTE DISORDER, TEARY EYES, TEMP, TEMP 36.8, TEMP 36.9, TEMPERATURE, TEMPERATURE 36.8, TEMPERATURE 36.9, TEMPERATURE:, TEMPERATURE: 100, TEMPERATURE: 100.0, TEMPERATURE: 100.1, TEMPERATURE: 100.2, TEMPERATURE: 100.3, TEMPERATURE: 100.4, TEMPERATURE: 100.5, TEMPERATURE: 100.6, TEMPERATURE: 100.7, TEMPERATURE: 100.8, TEMPERATURE: 100.9, TEMPERATURE: 101, TEMPERATURE: 101.0, TEMPERATURE: 101.1, TEMPERATURE: 101.2, TEMPERATURE: 101.3, TEMPERATURE: 101.4, TEMPERATURE: 101.5, TEMPERATURE: 101.6, TEMPERATURE: 101.7, TEMPERATURE: 101.8, TEMPERATURE: 101.9, TEMPERATURE: 102, TEMPERATURE: 102.0, TEMPERATURE: 102.5, TEMPERATURE: 103, TEMPERATURE: 103.0, TEMPERATURE: 104, TEMPERATURE: 35.3, TEMPERATURE: 35.4, TEMPERATURE: 35.5, TEMPERATURE: 35.6, TEMPERATURE: 35.7, TEMPERATURE: 35.8, TEMPERATURE: 35.9, TEMPERATURE: 36, TEMPERATURE: 36.0, TEMPERATURE: 36.1, TEMPERATURE: 36.2, TEMPERATURE: 36.3, TEMPERATURE: 36.4, TEMPERATURE: 36.5, TEMPERATURE: 36.6, TEMPERATURE: 36.7, TEMPERATURE: 36.8, TEMPERATURE: 36.9, TEMPERATURE: 37, TEMPERATURE: 37., TEMPERATURE: 37.0, TEMPERATURE: 37.1, TEMPERATURE: 37.2, TEMPERATURE: 37.3, TEMPERATURE: 37.4, TEMPERATURE: 37.5, TEMPERATURE: 37.6, TEMPERATURE: 37.7, TEMPERATURE: 37.8, TEMPERATURE: 37.9, TEMPERATURE: 38, TEMPERATURE: 38., TEMPERATURE: 38.0, TEMPERATURE: 38.1, TEMPERATURE: 38.2, TEMPERATURE: 38.3, TEMPERATURE: 38.4, TEMPERATURE: 38.5, TEMPERATURE: 38.6, TEMPERATURE: 38.7, TEMPERATURE: 38.8, TEMPERATURE: 38.9, TEMPERATURE: 39, TEMPERATURE: 39.0, TEMPERATURE: 39.1, TEMPERATURE: 39.2, TEMPERATURE: 39.3, TEMPERATURE: 39.4, TEMPERATURE: 39.5, TEMPERATURE: 39.6, TEMPERATURE: 39.7, TEMPERATURE: 39.8, TEMPERATURE: 39.9, TEMPERATURE: 40, TEMPERATURE: 40.0, TEMPERATURE: 40.1, TEMPERATURE: 98, TEMPERATURE: 99, TEMPERATURE: 99.0, TEMPERATURE: 99.1, TEMPERATURE: 99.2, TEMPERATURE: 99.4, TEMPERATURE: 99.5, TEMPERATURE: 99.6, TEMPERATURE: 99.7, TEMPERATURE: 99.8, TEMPERATURE: 99.9, TESTED POSITIVE ON RAPID TEST, THROAT, THROAT CONGESTION, THROAT IRRITATION, THROAT PAIN, THROAT TICKLE, THROWING UP, TICKLE IN THROAT, TICKLE THROAT, TIGHT CHEST, TIGHTNESS, TIGHTNESS IN CHEST, TIGHTNESS IN THE CHEST, TIGHTNESS OF CHEST, TIRED, TIRED HEADACHE, TIRED., TIREDNESS, TO BE ADMITTED, TODAY, TRANSFER, TRANSFER TO LTC, TRAUMA, TRAVEL, TRIEDNESS, TROUBLE BREATHING,

TROUBLE SWALLOWING, TUMMY ACHE, U, UK, UN, UN VACC, UNEXP FATIGUE, UNEXPLAINED FATIGUE, UNHOUSED, UNIMMUNIZED OR INCOMPLETE, UNK, UNKNOWN, UNKNOWN COUGH, UNKNOWN FEVER, UNKNOWN PNEUMONIA, UNKNOWN SOB, UNKNOWN SORE THROAT, UNPROTECTED SEX, UNVACCINATED, UNWELL, UPCOMING SX, UPSET STOMACH, UPSET STOMACH HEADACHE, UPSET STOMACHE, URINARY TRACT INFECTION, UTI, V, VACCINATED, VAGINAL BLEEDING, VAGINAL DISCHARGE, VAXXED, VERTIGO, VERY TIRED, VESICULAR RASH, VISIT LTC, VISITING LTC, VITALS STABLE., VOICE, VOICE CHANGE, VOICE LOSS, VOLUNTEER, VOM, VOMIT, VOMIT DIARRHEA, VOMIT X1, VOMITED, VOMITED X 1, VOMITED X1, VOMITIN, VOMITING, VOMITING AND DIARRHEA, VOMITING AND HEADACHE, VOMITING DIARRHEA, VOMITING HEADACHE, VOMITING NAUSEA, VOMITING OR DECREASED DRINKING, VOMITING RUNNY NOSE, VOMITING X1, VOMITING., VOMITING. DIARRHEA, VOMITINGS, VOMITNG, VOMITTED, VOMITTING, VOMITTING DIARRHEA, VOMITTING., VOMMIT, VOMMITING, VOMMITTING, VOMTING, VS NA, VSA, VSS, Vaginal bleeding, WARM, WARM TO TOUCH, WATERY EYES, WEAK, WEAKNESS, WEIGHT LOSS, WET COUGH, WHEEZE, WHEEZING, WHEEZY, WORK, WORK CLEARANCE, WORK REQUIREMENT, WORKS IN LTC, WORSENING COUGH, Y, YELLOW PHLEGM, YES, YES COUGH, YES FEVER, YES PNEUMONIA, YES SOB, YES SORE THROAT, YES- NOT SPECIFIED, YM

**Table 1.** Descriptive characteristics of children aged 6 months to 5 years tested for SARS-CoV-2 with COVID-19-related symptoms between July 28, 2022, and December 31, 2022 comparing unvaccinated with vaccinated (n=4,039)

| | Unvaccinated, n (%)[a] | Vaccinated, n (%)[a] | SD[b] |
|---|---|---|---|
| Total | 3772 | 267 | |
| Age group | | | |
| Age 6 months to <1 year | 958 (25.4) | 55 (20.6) | 0.26 |
| Age 1 year to <2 years | 774 (20.5) | 68 (25.5) | 0.11 |
| Age 2 to <3 years | 701 (18.6) | 63 (23.6) | 0.12 |
| Age 3 to <4 years | 522 (13.8) | 49 (18.4) | 0.12 |
| Age 4 to <5 years | 817 (21.7) | 32 (12.0) | 0.12 |
| Male sex | 2073 (55.0) | 151 (56.6) | 0.03 |
| Any comorbidity[c] | 195 (5.2) | 21 (7.9) | 0.11 |
| Past influenza vaccination | 811 (21.5) | 166 (62.2) | 0.91 |
| Mother healthcare worker status | 364 (9.7) | 72 (27.0) | 0.46 |
| Public health unit region[d] | | | |
| Central East | 189 (5.0) | 14 (5.2) | 0.01 |
| Central West | 673 (17.8) | 57 (21.3) | 0.09 |
| Durham | 69 (1.8) | 9 (3.4) | 0.10 |
| Eastern | 128 (3.4) | 14 (5.2) | 0.09 |
| Northern | 1295 (34.3) | 77 (28.8) | 0.12 |
| Ottawa | 52 (1.4) | 19 (7.1) | 0.29 |
| Peel | 356 (9.4) | 6 (2.2) | 0.31 |
| South West | 768 (20.4) | 33 (12.4) | 0.22 |
| Toronto | 153 (4.1) | 29 (10.9) | 0.26 |
| York | 68 (1.8) | 9 (3.4) | 0.01 |
| Area-level income quintile[d,e] | | | |
| 1 | 832 (22.1) | 28 (10.5) | 0.32 |
| 2 | 726 (19.2) | 35 (13.1) | 0.17 |
| 3 | 762 (20.2) | 63 (23.6) | 0.08 |
| 4 | 757 (20.1) | 68 (25.5) | 0.13 |
| 5 | 644 (17.1) | 72 (27.0) | 0.24 |
| Area-level household density quintile[d] | | | |
| 1 | 717 (19.0) | 43 (16.1) | 0.08 |
| 2 | 969 (25.7) | 65 (24.3) | 0.03 |
| 3 | 575 (15.2) | 47 (17.6) | 0.06 |
| 4 | 753 (20.0) | 59 (22.1) | 0.05 |
| 5 | 710 (18.8) | 52 (19.5) | 0.02 |
| Area-level visible minority quintile[d] | | | |
| 1 | 1225 (32.5) | 81 (30.3) | 0.05 |
| 2 | 894 (23.7) | 65 (24.3) | 0.02 |
| 3 | 565 (15.0) | 58 (21.7) | 0.18 |

| | | | |
|---|---|---|---|
| 4 | 491 (13.0) | 43 (16.1) | 0.09 |
| 5 | 549 (14.6) | 20 (7.5) | 0.23 |
| Essential workers quintile[d,f] | | | |
| 1 | 331 (8.8) | 59 (22.1) | 0.38 |
| 2 | 841 (22.3) | 65 (24.3) | 0.05 |
| 3 | 836 (22.2) | 70 (26.2) | 0.10 |
| 4 | 863 (22.9) | 50 (18.7) | 0.10 |
| 5 | 853 (22.6) | 23 (8.6) | 0.39 |
| Hospitalization related to COVID-19 (%) | 216 (5.7) | 6 (2.2) | 0.18 |

[a]Proportion reported, unless stated otherwise.
[b]SD=standardized difference. Standardized differences of >0.15 are considered clinically relevant. Comparing unvaccinated subjects to vaccinated subjects.
[c]Comorbidities include asthma, diabetes, immunocompromising conditions caused by underlying diseases or therapy, autoimmune diseases, active cancer, or pediatric complex chronic conditions.
[d]The sum of counts does not equal the column total because of individuals with missing information (≤2.0%) for this characteristic.
[e]Household income quintile has variable cut-off values in each city or Census area to account for cost of living. A dissemination area (DA) being in quintile 1 means it is among the lowest 20% of DAs in its city by income.
[f]Percentage of people in the area working in the following occupations: sales and service occupations; trades, transport and equipment operators and related occupations; natural resources, agriculture, and related production occupations; and occupations in manufacturing and utilities. Census counts for people are randomly rounded up or down to the nearest number divisible by 5, which causes some minor imprecision.

**Figure 1.** Vaccine effectiveness estimates in children aged 6 months to 5 years old against symptomatic SARS-CoV-2 infection, by month after initial dose

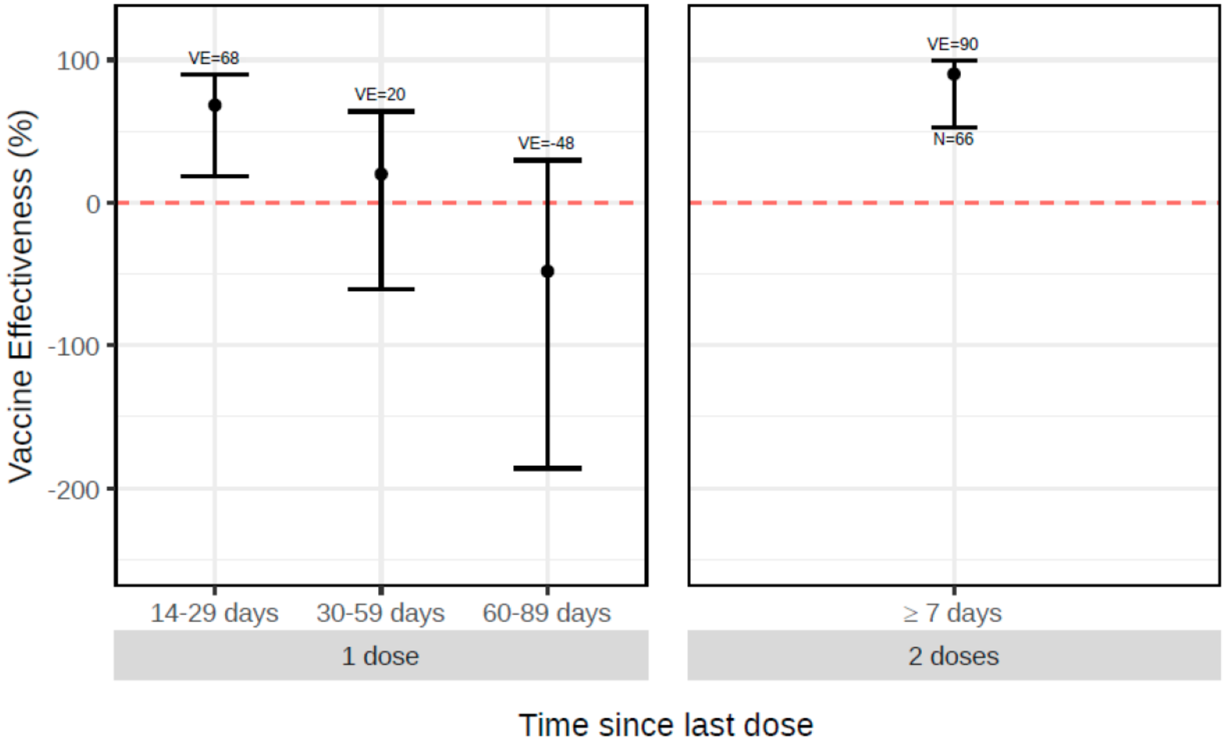

**Table 2.** Comparison of unadjusted and adjusted VE estimates against symptomatic SARS-CoV-2 infection, adjusted for a single covariate to evaluate the impact of potential confounders

| | ≥14 days after dose 1 | | | | ≥7 days after dose 2 | | | |
|---|---|---|---|---|---|---|---|---|
| | VE | Lower 95% CI | Upper 95% CI | %change in OR[a] | VE | Lower 95% CI | Upper 95% CI | %change in OR[a] |
| Unadjusted | 0.12 | -0.32 | 0.44 | 0.00 | 0.91 | 0.59 | 0.99 | 0.00 |
| Age group | 0.04 | -0.45 | 0.38 | 9.60 | 0.91 | 0.58 | 0.99 | 1.34 |
| Sex | 0.12 | -0.32 | 0.44 | 0.08 | 0.91 | 0.59 | 0.99 | 0.05 |
| Any comorbidity | 0.12 | -0.31 | 0.44 | -0.17 | 0.91 | 0.59 | 0.99 | -0.29 |
| Past influenza vaccine | 0.12 | -0.33 | 0.44 | 0.24 | 0.91 | 0.58 | 0.99 | 0.28 |
| HCW mother | 0.11 | -0.34 | 0.43 | 1.66 | 0.91 | 0.58 | 0.99 | 1.31 |
| Public health unit | 0.29 | -0.09 | 0.55 | -18.94 | 0.93 | 0.67 | 1.00 | -21.76 |
| Income quintile | 0.13 | -0.30 | 0.44 | -1.26 | 0.91 | 0.59 | 0.99 | -1.10 |
| Household density quintile | 0.12 | -0.32 | 0.43 | 0.21 | 0.91 | 0.60 | 0.99 | -2.71 |
| Visible minority quintile | 0.09 | -0.37 | 0.42 | 3.48 | 0.91 | 0.57 | 0.99 | 3.47 |
| Essential worker quintile | 0.22 | -0.17 | 0.50 | -11.59 | 0.92 | 0.63 | 1.00 | -11.08 |
| Week of test | 0.07 | -0.42 | 0.41 | 6.20 | 0.87 | 0.41 | 0.99 | 40.49 |
| Number of physician visits | 0.10 | -0.35 | 0.42 | 2.62 | 0.92 | 0.63 | 1.00 | -12.12 |
| Previous infection | 0.09 | -0.36 | 0.42 | 3.16 | 0.91 | 0.60 | 0.99 | -2.04 |

[a]% Change in OR = (adjusted OR/crude OR - 1)*100
